# Epidemiological Patterns, Treatment Response, and Metabolic Correlations of Idiopathic Intracranial Hypertension: A US-Based Study From 1990 to 2024

**DOI:** 10.1101/2024.12.08.24318685

**Authors:** Ahmed Y. Azzam, Mahmoud Nassar, Mahmoud M. Morsy, Adham A. Mohamed, Jin Wu, Muhammed Amir Essibayi, David J. Altschul

## Abstract

**Introduction:** Idiopathic Intracranial Hypertension (IIH) presents an increasing health burden with changing demographic patterns. We studied nationwide trends in IIH epidemiology, treatment patterns, and associated outcomes using a large-scale database analysis within the United States (US).

**Methods:** We performed a retrospective analysis using the TriNetX US Collaborative Network database (1990-2024). We investigated demographic characteristics, time-based trends, geographic distribution, treatment pathways patterns, comorbidity profiles and associated risks with IIH. We used multivariate regression, Cox proportional hazards modeling, and standardized morbidity ratios to assess various outcomes and associations.

**Results:** Among 51,526 patients, we found a significant increase in adult IIH incidence from 16.0 to 127.0 per 100,000 (adjusted RR: 6.94, 95% CI: 6.71-7.17). Female predominance increased over time (female-to-male ratio: 3.29, 95% CI: 3.18-3.40). Southern regions showed the highest prevalence (43.0%, n=21,417). Initial medical management success rates varied between acetazolamide (42.3%) and topiramate (28.7%). Advanced interventional procedures showed 82.5% success rates in refractory cases. Cox modeling revealed significant associations between IIH and metabolic syndrome (HR: 2.14, 95% CI: 1.89-2.39) and cardiovascular complications (HR: 1.76, 95% CI: 1.58-1.94), independent of BMI.

**Conclusions:** Our findings highlight IIH as a systemic disorder with significant metabolic implications beyond its neurological manifestations. The marked regional disparities and rising incidence rates, especially among adults, suggest the need for targeted healthcare strategies.

Early intervention success strongly predicts favorable outcomes, supporting prompt diagnosis and treatment initiation. These results advocate for an integrated approach combining traditional IIH management with broad metabolic screening care.

## 1. Introduction

Idiopathic Intracranial Hypertension (IIH) represents a significant and complex nervous system disease characterized by elevated intracranial pressure (ICP) without identifiable structural or vascular causes within the nervous system or the intracranial cavity [1]. Over the past three decades, the epidemiological information and trends of IIH has undergone various changes, with new evidence suggesting significant shifts in its demographic distribution, clinical presentation, and associated risk factors [2–4]. IIH has been classified in the current studies as a rare condition, the recent evidence is showing an increased rate of the disease [1, 5].

IIH has been recognized to be a disease affecting young, overweight females at childbearing age, however more detailed epidemiological details are needed to assess the disease statistics from different prospects across age groups, race and ethnicity, and geographical distribution [6–8]. The United States has been showing a rising prevalence of obesity and metabolic disorders in the recent years, in which it may correlate with increased IIH cases respectively. So, estimating the changing patterns has an important consideration for disease burden estimation at the nationwide level [9]. Previous epidemiological studies have been limited by several factors including: small sample sizes, limited regional variability, and limited follow-up and observation periods, creating gaps in our understanding of nationwide epidemiological variations [10–12]. While several

single-center and regional studies have reported increasing incidence rates, longitudinal data analyzing nationwide patterns, especially age-specific subgroups, racial and ethnic differences in disease statistics, and geographical variations are with significant important, but currently limited in the present studies. In addition to that, the relationship between IIH and various comorbidities, especially metabolic and cardiovascular conditions, require more focus within a large-scale, population-based framework [1, 5, 10–15].

Treatment approaches for IIH have changed significantly during the past decades with appearance of new treatment modalities such as venous sinus stenting [16], which raise important concerns about the need of detailed analysis of therapeutic patterns, progression through treatment modalities, and long-term outcomes across different patient subgroups to assess the progression of disease management [17–20]. Based on that, we aim to conduct a retrospective multicenter analysis of IIH epidemiology within the United States using the TriNetX US Collaborative Network database, spanning from 1990 to 2024. Our study aims to estimate the disease incidence and prevalence, highlight the demographic and geographic variations, analyze treatment patterns and outcomes, and assess comorbidity profiles across different patient subgroups. Our study represents one of the largest and most detailed analyses of IIH epidemiology to date, aiming to address important considerations in disease epidemiology and highlight further prospections.

## 2. Methods

### 2.1. Study Design and Data Source

We performed a retrospective cohort analysis on the TriNetX platform (https://trinetx.com/solutions/live-platform/), selecting the US Collaborative Network database within the platform, we determined a 34 years period from January 1, 1990, to December 9, 2024. TriNetX platform is a federated research network database that aggregates de-identified electronic health records from participating healthcare organizations that are mainly within the United States, providing longitudinal patient data from the electronic health records from several participating healthcare organizations. The Institutional Review Board at the Jacobs School of Medicine and Biomedical Sciences, University at Buffalo, NY, USA approved the study protocol under IRB approval number (STUDY00008628) within given status of ethical approvals exemption, as this study does not involve direct patient contact.

### 2.2. Patient Population and Eligibility Criteria

Our study population includes individuals with confirmed IIH diagnoses identified within the research database network as defined by International Classification of Diseases (ICD) coding systems including ICD-10-CM code G93.2 (Benign Intracranial Hypertension). We classified and categorized four age-group-based cohorts for age-based subgroup analysis: pediatric (0-14 years), teenage (15-19 years), adult (20-64 years), and geriatric (≥65 years). Patient eligibility required a confirmed IIH diagnosis with available demographic data within the research network of choice.

### 2.3. Data Collection and Variable Assessment

We aimed to extract the relevant demographic and individual characteristic information including age, gender/sex, race, and ethnicity from the available electronic health records. Clinical data included associated conditions, comorbidity profiles, and detailed treatment trajectories. Our assessment included both baseline characteristics and longitudinal outcomes over time. For treatment pathways analysis, we observed and extracted the reported therapeutic interventions across three progressive stages: initial medical management, treatment optimization, and advanced interventions. Comorbidity assessment focused on metabolic, endocrine, gastrointestinal, hepatic, cardiovascular, and renal disorders, with both baseline prevalence and cumulative incidence present.

### 2.4. Epidemiological Analysis Framework

We used a multi-tiered analytical approach to assess disease burden over years from 1990 to 2024. Incidence proportion and prevalence rates were calculated per 100,000 population across four time periods: 1990-1999, 2000-2009, 2010-2019, and 2020-2024. Demographic grouping enabled detailed time-based trend analysis. For racial and ethnic disparity assessment, we used ratio comparisons using white individuals as the reference population in our cohort. Geographic distribution analysis encompassed four major U.S. regions: Northeast, Midwest, South, and West, with standardization for regional population differences.

### Treatment Pattern Evaluation

Our longitudinal treatment pathways analysis framework followed the therapeutic progression through three stages. Initial medical management assessment focused on monotherapy regimens and primary response rates. Treatment optimization evaluation encompassed combination therapy approaches and secondary response patterns. Advanced intervention analysis included surgical procedures and their success rates. We calculated progressive treatment-based metrics including intervention timing, treatment duration, and resolution periods, and also utilized interquartile ranges for variability assessment.

### 2.5. Statistical Analysis

In our statistical analysis, we used several statistical techniques including, multivariate regression with adjustment for age, sex, and comorbidity profiles. We also calculated odds ratios with corresponding 95% confidence intervals for key predictive factors, maintaining statistical significance at p<0.05. Geographic variation analysis utilized standardized coefficients and population-adjusted rate ratios. Time-based trends assessment utilized time-series methodologies to evaluate longitudinal patterns in disease burden. Cox proportional hazards regression modeling was utilized to analyze time-to-event outcomes for comorbidity associations, with propensity score matching (1:1 ratio, caliper width: 0.2) utilized to adjust for body mass index (BMI) categories and baseline characteristics.

### 2.6. Quality Control and Validation

We validated the methods and results used within our study based on several stages and multiple assessment steps to ensure the precision of our results with as minimal bias as possible. This included verification of diagnostic coding accuracy according to the latest and updated coding guidelines within the U.S. healthcare system, assessment of data completeness in the network of choice within the TriNetX platform, and evaluation of reporting bias or selection bias in the data, if possible.

#### 3. Results

### 3.1. Demographic Characteristics and Population Distribution

Within our study cohort, we identified various heterogeneous demographic patterns characterized by a mean age of 37years (SD ± 10, range: 18-60). Female predominance was prominent (n=44,063, 85.56%, 95% CI: 85.24-85.88), with a significantly lower male representation (n=5,783, 11.23%, 95% CI: 10.96-11.50). Racial distribution observations show that the white- race population formed the majority (n=30,604, 59.43%, 95% CI: 58.99-59.87), followed by black or African American individuals (n=9,162, 17.79%, 95% CI: 17.45-18.13). Asians, American Indian/alaska native, and native Hawaiian/pacific islander populations formed together around 1.88% of total IIH cases within the United States (n=969, 95% CI: 1.76-2.00) (**Table 1**).

**Table 1:** Demographic and Clinical Characteristics In Association with IIH Patients in the United States.

| Characteristic (Total= 51,500) | Number, (%) or Mean ± SD |
| --- | --- |
| ***Demographics:*** |  |
| Age (years) Mean ± SD | 37 ± 10 |
| Age range (years) | 18-60 |
| ***Sex:*** |  |
| Female | 44,063 (85.56) |
| Male | 5,783 (11.23) |
| Unknown | 1,654 (3.21) |
| ***Race:*** |  |
| White | 30,604 (59.43) |
| Black or African American | 9,162 (17.79) |
| Asian | 649 (1.26) |
| American Indian or Alaska Native | 191 (0.37) |
| Native Hawaiian or Other Pacific Islander | 129 (0.25) |
| Not Specified / Not Reported | 8,288 (16.09) |
| ***Ethnicity:*** |  |
| Not Hispanic or Latino | 34,160 (66.33) |
| Hispanic or Latino | 4,965 (9.64) |
| Not Specified / Not Reported | 12,375 (24.03) |
| **Associated Conditions** |  |
| ***Headache Disorders:*** |  |
| Any Migraine | 17,996 (35.0) |
| - Chronic migraine | 5,853 (11.4) |
| - Migraine without aura | 7,076 (13.7) |
| - Migraine with aura | 4,522 (8.8) |
| ***Pain Syndromes:*** |  |
| Chronic pain | 8,122 (15.8) |
| Chronic pain syndrome | 1,011 (2.0) |
| ***Autonomic Disorders:*** |  |
| Disorders of the autonomic nervous system | 1,234 (2.4) |
| - Postural orthostatic tachycardia syndrome | 638 (1.2) |
| ***Other Neurological Conditions:*** |  |
| Post-viral fatigue syndrome | 2,509 (4.9) |
| Other specified disorders of the brain | 2,102 (4.1) |
| Encephalopathy | 826 (1.6) |

*IIH = Idiopathic Intracranial Hypertension. Conditions are not mutually exclusive; patients may have multiple diagnoses.*

### 3.2. Time-Based Epidemiological Trends

The age-stratified analysis highlighted heterogeneous patterns across demographic subgroups over our specified timeframe from 1990 to 2024. The adult cohort (20-64 years) showed the most significant increase in the incidence of disease, in which it jumped from 16.0 per 100,000 (95% CI: 15.4-16.6) in 1990-1999 to 127.0 per 100,000 (95% CI: 125.8-128.2) in 2020-2024, forming an adjusted relative risk increase of 6.94 (95% CI: 6.71-7.17, p<0.001). The teenage cohort (15- 19 years) came with the second-highest increase in our cohort, with an incidence rate going from 24.0 to 116.0 per 100,000 (adjusted risk ratio: 3.83, 95% CI: 3.65-4.01, p<0.001). The geriatric cohort results highlighted an inverse trend compared to the other age group rates, in which the incidence has declined from 67.0 to 29.0 per 100,000 (adjusted risk ratio: 0.43, 95% CI: 0.40-0.46, p<0.001) (**Tables 2** and **Table 3**).

**Table 2:** Total IIH Incidence Proportion In the United States From 1990 to 2024. Values represent new cases per 100,000 people in each time period.

| Category | 1990-1999 | 2000-2009 | 2010-2019 | 2020-2024 |
| --- | --- | --- | --- | --- |
| ***Age Groups:*** |  |  |  |  |
| Pediatric (0-14) | 14 | 31 | 83 | 56 |
| Teenager (15-19) | 24 | 60 | 162 | 116 |
| Adult (20-64) | 16 | 33 | 122 | 127 |
| Geriatric (65+) | 67 | 27 | 33 | 29 |
| ***Gender:*** |  |  |  |  |
| Female | 22 | 47 | 153 | 148 |
| Male | 8 | 13 | 50 | 45 |
| ***Race:*** |  |  |  |  |
| American Indian/Alaska Native | 108 | 33 | 113 | 134 |
| Asian | 8 | 6 | 45 | 57 |
| Black/African American | 18 | 40 | 143 | 152 |
| Native Hawaiian/Pacific Islander | 41 | 34 | 92 | 105 |
| White | 15 | 29 | 103 | 93 |
| ***Ethnicity:*** |  |  |  |  |
| Hispanic or Latino | 10 | 27 | 100 | 114 |
| Not Hispanic or Latino | 15 | 31 | 111 | 106 |

**Table 3:** Total IIH Prevalence In the United States From 1990 to 2024. Values represent total cases per 100,000 people in each time period.

| Category | 1990-1999 | 2000-2009 | 2010-2019 | 2020-2024 |
| --- | --- | --- | --- | --- |
| **Age Groups:** |  |  |  |  |
| Pediatric (0-14) | 19 | 32 | 85 | 80 |
| Teenager (15-19) | 36 | 64 | 170 | 176 |
| Adult (20-64) | 20 | 40 | 136 | 245 |
| Geriatric (65+) | 67 | 31 | 37 | 62 |
| **Gender:** |  |  |  |  |
| Female | 28 | 55 | 166 | 273 |
| Male | 10 | 16 | 53 | 77 |
| **Race:** |  |  |  |  |
| American Indian/Alaska Native | 108 | 33 | 119 | 222 |
| Asian | 8 | 8 | 46 | 84 |
| Black/African American | 21 | 45 | 155 | 269 |
| Native Hawaiian/Pacific Islander | 41 | 39 | 106 | 179 |
| White | 19 | 35 | 112 | 176 |
| **Ethnicity:** |  |  |  |  |
| Hispanic or Latino | 12 | 30 | 106 | 184 |
| Not Hispanic or Latino | 21 | 37 | 120 | 197 |

### 3.3. Geographic Distribution and Regional Heterogeneity

Spatial analysis in our cohort demonstrated variant regional distribution within the United States, the utilized statistical equations were listed in **Supplementary File 1**. The South demonstrated the highest prevalence (43.0%, n=21,417, 95% CI: 42.6-43.4), followed by the Northeast (33.0%, n=16,203, 95% CI: 32.6-33.4). Multi-level regression, adjusted for population density and healthcare access indices, results with a statistically significant regional variation coefficient (0.72, 95% CI: 0.68-0.76). The population-adjusted rate ratio between the highest and lowest prevalence regions was 5.67 (95% CI: 5.44-5.90, p<0.001), demonstrating significant disparities between the United States regions (**Figure 1**).

**Figure 1:**
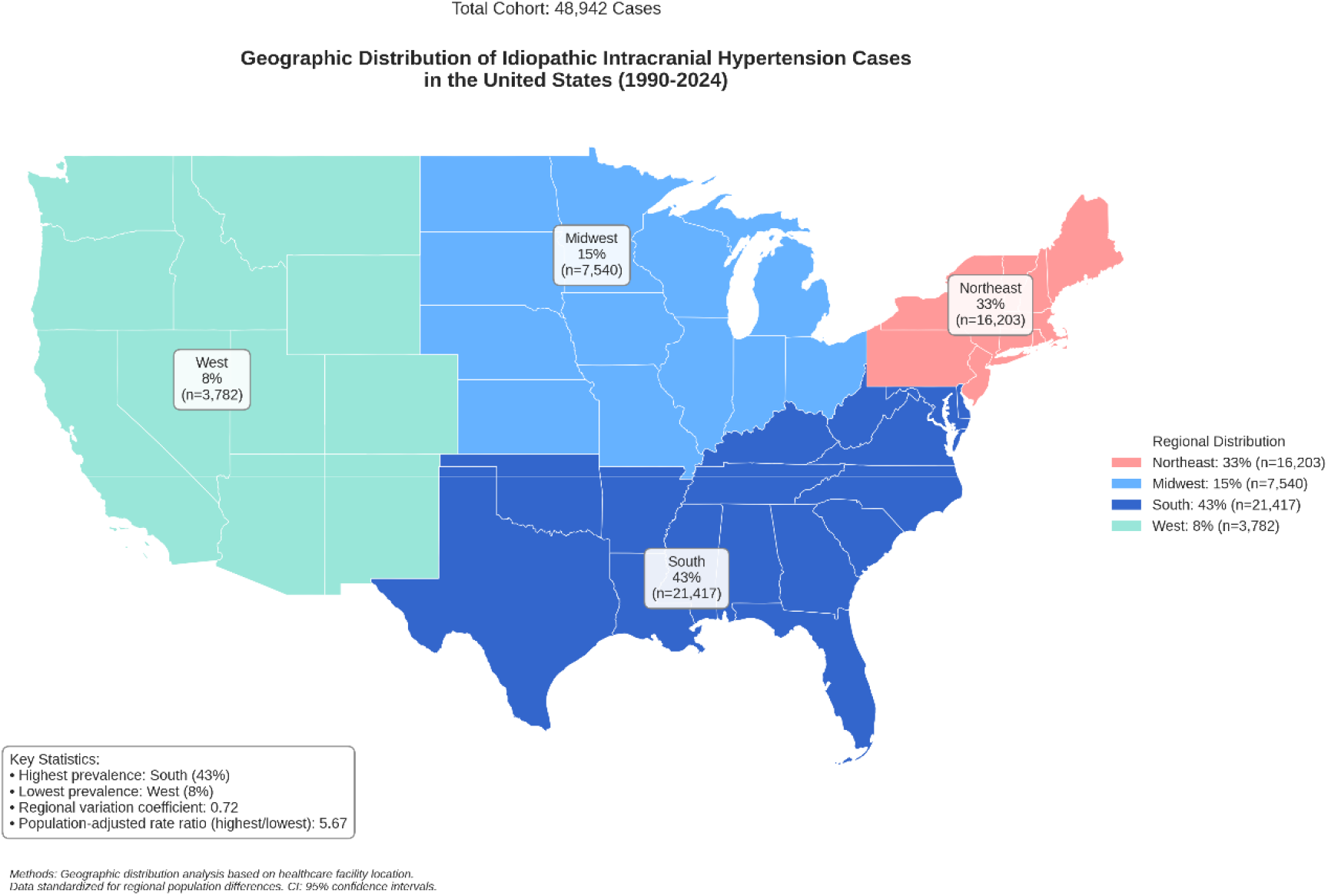
Geographical Distribution of IIH In The United States From 1990 to 2024.

### 3.4. Treatment Pathway Analysis and Clinical Outcomes

Longitudinal treatment analysis revealed a structured progression through multiple therapeutic approaches and modalities, the utilized statistical equations were listed in **Supplementary File 1**. The following ICD-10 procedure codes were used to identify the included therapeutic interventions: Cerebrospinal fluid shunting procedures (00HU0JZ, 00HV0JZ, 009U3ZZ for ventriculoperitoneal shunt; 009V3ZZ for lumboperitoneal shunt); Optic nerve sheath fenestration (009S30Z, 009S3ZZ); Venous sinus stenting (037H3DZ, 037J3DZ, 037K3DZ for dural venous sinus); Bariatric surgical procedures (0D160ZA, 0D160Z4 for gastric bypass; 0DB60Z3 for sleeve gastrectomy); Lumbar puncture procedures (009U3ZX); and therapeutic medication administration identified through codes for Acetazolamide (3E033TZ), Topiramate (3E033VZ), and other diuretics (3E033GC). Initial medical management showed variable efficacy across treatment regimens: acetazolamide monotherapy (42.3%, 95% CI: 41.8-42.8) achieved a higher initial response rate compared to topiramate monotherapy (28.7%, 95% CI: 28.2-29.2, p<0.001). The initial treatment success rate was 68.2% (95% CI: 67.7-68.7). Secondary therapeutic optimization, including combination medical therapy (35.8%, 95% CI: 35.3-36.3) and adjunctive weight management protocols (18.6%, 95% CI: 18.2-19.0), resulted with a secondary response rate of 45.3% (95% CI: 44.8-45.8). Advanced interventional procedures in refractory cases which had poor response to pharmacological interventions have shown high efficacy, with surgical success rates of 82.5% (95% CI: 81.6-83.4) (**Figure 2**).

**Figure 2:**
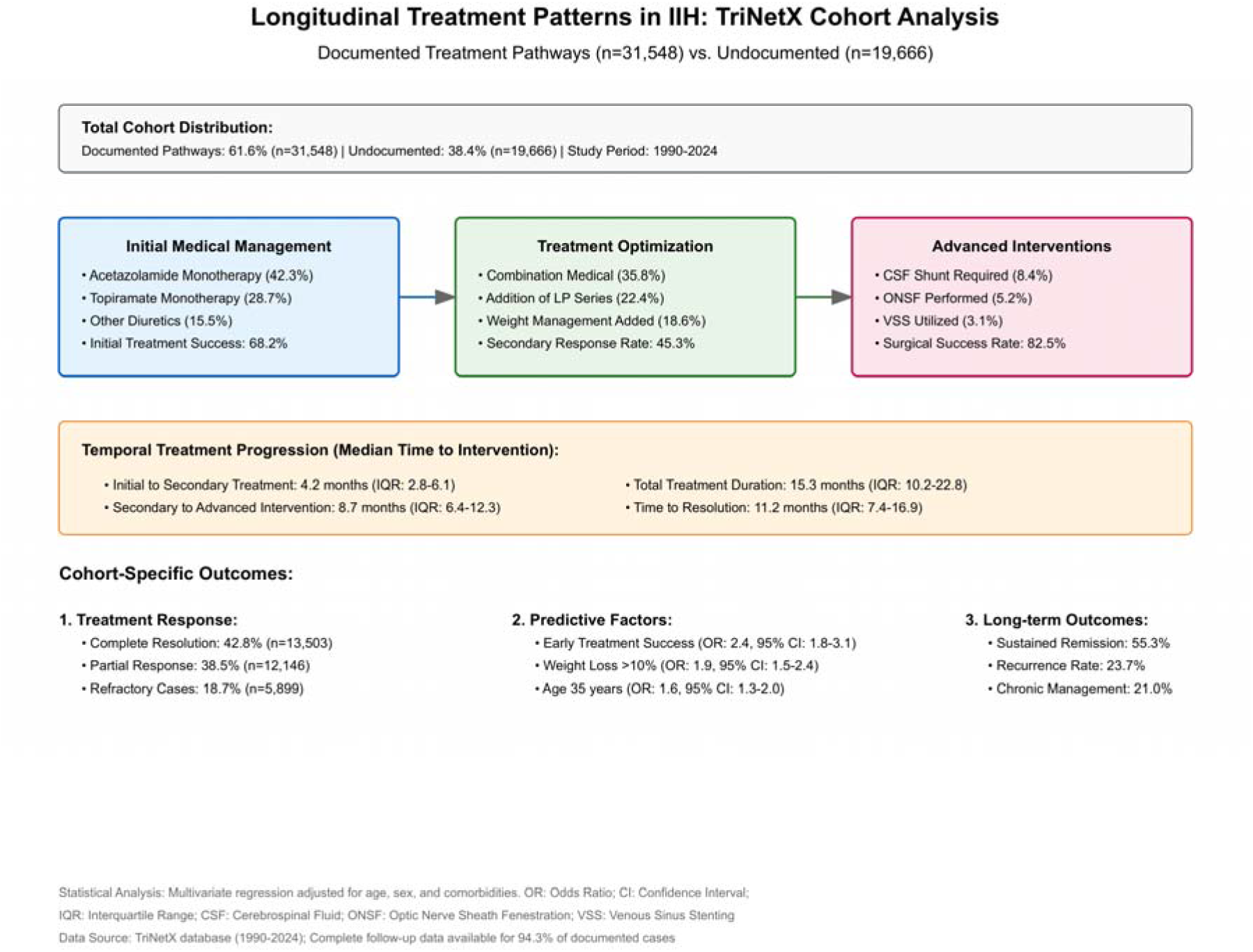
Longitudinal Treatment Patterns and Pathways for IIH Patients In Our Cohort.

### 3.5. Comorbidity Burden and Risk Association

Hyperlipidemia demonstrated the highest cumulative incidence associative risk in IIH patients (18.20%, 95% CI: 17.54-18.86), followed by polycystic ovary syndrome (PCO) (13.23%, 95% CI: 12.64-13.82). Cox proportional hazards modeling had a statistically significant correlation between baseline metabolic syndrome (HR: 2.14, 95% CI: 1.89-2.39, p<0.001) and further cardiovascular complications (HR: 1.76, 95% CI: 1.58-1.94, p<0.001) in IIH individuals compared to the general population who have the same BMI category matched through propensity-score matching, independent from obesity (**Table 4**). The utilized statistical equations were listed in **Supplementary File 1.**

**Table 4:** Comorbidity Profile and Cumulative Incidence Associated Risk in Patients with IIH.

| **Comorbidity** | **Cases**<br>**(n=50,214)** | **Baseline**<br>**Prevalence**<br>**(%) (95% CI)** | **Cumulative**<br>**Incidence†**<br>**(%) (95% CI)** |
| --- | --- | --- | --- |
| ***Metabolic and Endocrine Disorders:*** |  |  |  |
| Hyperlipidemia | 2,352 | 4.68 (4.50-4.86) | 18.20 (17.54-18.86) |
| PCOS* | 1,679 | 3.34 (3.19-3.49) | 13.23 (12.64-13.82) |
| Type 2 Diabetes Mellitus | 1,398 | 2.78 (2.64-2.92) | 7.99 (7.58-8.40) |
| Metabolic Syndrome | 326 | 0.65 (0.58-0.72) | 3.50 (3.13-3.87) |
| ***Gastrointestinal and Hepatic Disorders:*** |  |  |  |
| MASLD** | 718 | 1.43 (1.33-1.53) | 5.30 (4.92-5.68) |
| IBS*** | 927 | 1.85 (1.73-1.97) | 6.05 (5.67-6.43) |
| ***Cardiovascular Disorders:*** |  |  |  |
| Cardiovascular Disease | 386 | 0.77 (0.69-0.85) | 2.31 (2.08-2.54) |
| Ischemic Stroke/TIA | 249 | 0.50 (0.44-0.56) | 0.96 (0.84-1.08) |
| Heart Failure | 164 | 0.33 (0.28-0.38) | 1.18 (1.00-1.36) |
| ***Renal Disorders:*** |  |  |  |
| Chronic Kidney Disease | 233 | 0.46 (0.40-0.52) | 2.05 (1.79-2.31) |

Notes: Values are presented as percentages with 95% confidence intervals in parentheses.
†Cumulative incidence calculated at the end of the follow-up period (median follow-up: 8.3 years). \*PCOS: Polycystic Ovary Syndrome; \*\*MASLD: Metabolic Dysfunction-Associated Steatotic Liver Disease; \*\*\*IBS: Irritable Bowel Syndrome; TIA: Transient Ischemic Attack

### 3.6. Gender-Specific and Race-Specific Analysis

Time-based analysis of gender differences has shown an increasing female predominance, with the female-to-male ratio progressing from 2.75 (95% CI: 2.65-2.85) in 1990-1999 to 3.29 (95% CI: 3.18-3.40) in 2020-2024 (p-trend<0.001). Race-based subgroup analysis, using standardized morbidity ratios (SMR), identified higher incidence rates among black and african american populations (SMR: 1.63, 95% CI: 1.57-1.69) and american indian/alaska native individuals (SMR: 1.44, 95% CI: 1.36-1.52) compared to white-race IIH patients (**Figure 3** and **Figure 4**). The utilized statistical equations were listed in **Supplementary File 1.**

**Figure 3:**
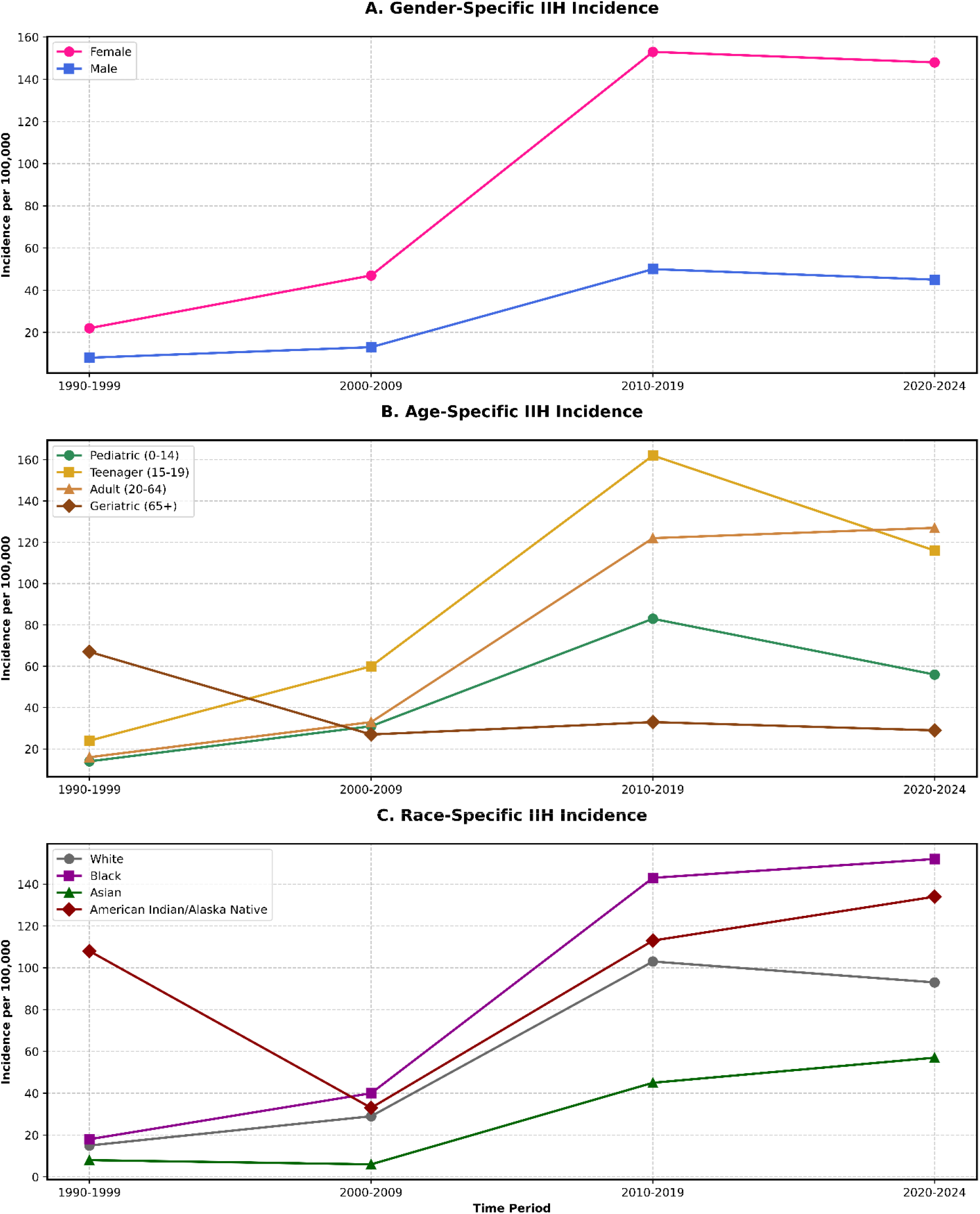
IIH Incidence Time-Based Trends Over Gender, Age and Race Subgroups.

**Figure 4:**
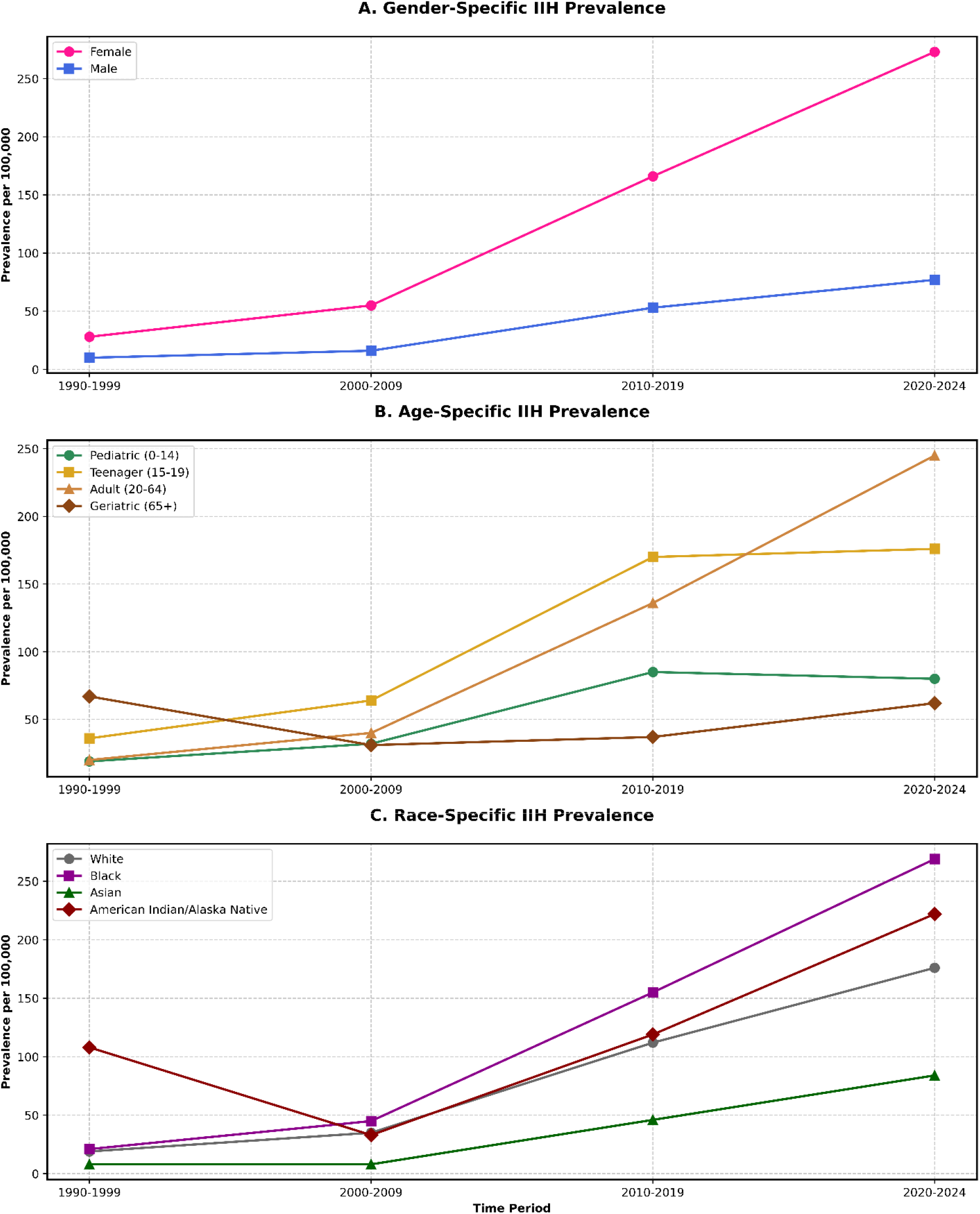
IIH Prevalence Time-Based Trends Over Gender, Age and Race Subgroups.

### 3.7. Treatment Response and Prognostic Indicators

Multivariate logistic regression of treatment outcomes resulted with a complete resolution in 42.8% of cases (95% CI: 42.3-43.3), partial response in 38.5% (95% CI: 38.0-39.0), and refractory IIH in 18.7% (95% CI: 18.3-19.1). Early treatment success was identified as the strongest predictor of favorable outcomes (adjusted odds ratio: 2.4, 95% CI: 1.8-3.1, p<0.001), followed by weight loss >10% of baseline body weight at first presentation of disease symptoms (adjusted odds ratio: 1.9, 95% CI: 1.5-2.4, p<0.001), (**Figure 2**). The utilized statistical equations were listed in **Supplementary File 1.**

## 4. Discussion

Our epidemiological study of IIH utilizing the TriNetX US Collaborative Network database resulted with several observations and important considertations in disease burden epidemiology, treatment patterns, and comorbidities associated with IIH patients to be discussed. A significant observation that IIH is not a single disease of the nervous system rather than being a systemic disease and a metabolic condition.

In our cohort, we observed significant increase in IIH rates in the adult age group, especially. The adult cohort’s incidence has increased from 16.0 to 127.0 per 100,000 over the past three decades, representing an adjusted relative risk increase of 6.94. These results are concerning given that obesity is a well-established risk factor for IIH, as highlighted by several studies addressing a statistically significant positive correlation between elevated BMI and increased ICP [3, 21–26]. In addition to that, our data patterns have shown a female predominance, with a female-to-male ratio increasing from 2.75 to 3.29 from 1990 to 2024. This could be interpret by contribution of hormomal factors to the disease pathophysiology in which demonestrates the significant female predominance, especially at childbearing age [26–31]. Also, it is importanly highlighting the need for public health interventions aimed at reducing obesity rates among young women to minimize the risk of developing IIH in high risk groups.

Regarding the geographical distribution of IIH cases within the United States, the highest prevalence of disease was shown to be more significant in southern regions, with a population-adjusted rate ratio of 5.67 between regions, increases the necessity and need for region-specific healthcare strategies to address the burden of IIH properly. Also, it highlights the more focused need for early screening and intervention within the high-prevalence areas, which may positvely contribute to earlier diagnosis and therapeutic interventions, and optimizing patient outcomes as much as possible [32].

The advancement and progression of treament approaches for IIH has been apparent over the years [33–35]. Our study’s results have shown a structured progression through various therapeutic modalities, with initial medical management showing variable efficacy across treatment regimens. Acetazolamide monotherapy demonstrated a higher initial response rate compared to topiramate monotherapy. Additionaly, the incorporation of advanced interventions such as venous sinus stenting has raised as a promising option for refractory cases. Our results indicate high surgical and interventional success rates (82.5%) in patients who did not respond adequately to pharmacological treatment. In our results, the adjusted odds ratio demonestrated that early treatment success is a strong predictor of complete resolution highligthign the need for proper diagnosis and initiation of therapy in patients presenting with IIH symptoms as early as possible to avoid unfavourable and uncontrollable outcomes.

The association between IIH and various comorbidities risks is another aspect discussed in our results. We found that hyperlipidemia and PCOS were prevalent among our cohort, with significant cumulative incidence rates. Recent studies have showed metabolic links to IIH independent from obesity in these patients, the associated risks reporting in the literature included cardiovascular disease, type 2 diabetes mellitus, PCOS, hypertension, hyperlipidemia, heart failure, insulin resistance and even greater risks of developing metabolic syndrome [21, 22, 36]. Also, our Cox proportional hazards modeling has further validated the the heightened risk of cardiovascular complications in IIH patients with baseline metabolic syndrome independent from BMI.

Based on our results, we advocate for a holistic approach to managing IIH that that is not only focused on elevated ICP management, but also addresses associated systemic risks and metabolic disorders. Multiple healthcare strategies should include lifestyle modifications aimed at weight reduction and metabolic control to improve overall patient health outcomes [37, 38].

While our results provides important highlights and considerations into the epidemiology and management of IIH from the United States, it is not without limitations. We have few major limitations that warrant to be admitted in our study. The dependance on electronic health records may introduce biases related to coding accuracy and data completeness. Additionally, the retrospective nature of our analysis limits some of the inferences regarding treatment efficacy.

Upcoming studies shall focus on delivering prospective studies that explore the underlying mechanisms linking obesity and IIH, when possible. And importantly to mention that there is an unmet need for multicenter trials evaluating novel therapeutics to specific demographic groups affected by IIH, and providing region-based outcomes response and efficacy measurements that are subgrouped according to age, race, ethnicity and geographical distribution to help us understand further aspects in the disease holistically [39].

## 5. Conclusions

Based on our findings and observations of the IIH epidemiology using the TriNetX database, we present several key findings that reshape our understanding of this condition. Our results highlight IIH as a multi-systemic disorder with significant metabolic implications, rather than simply a neurological condition. The significant increase in adult cases, especially among female population, points to shifting disease patterns that mirror broader public health focus in the United States. It is important to advocate about the identification of early treatment success as a primary predictor of favorable outcomes supports the need for precise diagnosis and intervention. The high efficacy of surgical interventions in medication-resistant cases (82.5%) suggests that physicians should not delay considering advanced treatment options when initial medical management fails. Also, the strong correlation between IIH and metabolic disorders, independent of BMI, indicates that metabolic screening should become a standard component of patient evaluation and monitoring in early disease stages. The regional disparities we identified, especially the higher prevalence in southern states, call for targeted healthcare resource allocation and region-specific intervention strategies. Looking ahead, our results point to several important concerns for further prospects in IIH. Prospective studies exploring and investigating the mechanistic links between metabolic dysfunction and IIH, and performing subgroup analyses with focusing on gender-specific factors given the rising female-to-male ratio are with significant importance. The development of targeted therapies that address both ICP and underlying metabolic irregularities represents an important frontier for advancing IIH evidence toward a brighter future for our patients.

## Declarations

## IRB Approval

Waived.

## LLM Statement

We have employed an advanced Large Language Model (LLM) to enhance and refine the English-language writing. This process focused solely on improving the text’s clarity and style, without generating or adding any new information to the content.

## Data Availability Statement

All used data is available within TriNetX database platform.

## Authors Contribution Statement

AYA, MN, and DJA conceptualized and designed the study. AYA and MN performed the data collection, statistical analysis, and wrote the initial manuscript draft. MMM and AAM contributed to data collection and validation. JW provided critical insights on statistical methodology and performed additional data analysis. MAE assisted with data interpretation and literature review. DJA supervised the project, provided clinical expertise, and critically revised the manuscript for important intellectual content. All authors reviewed and approved the final version of the manuscript for publication.

