## Supplementary File 1 for "Epidemiological Patterns, Treatment Response, and Metabolic Correlations of Idiopathic Intracranial Hypertension: A US-Based Study From 1990 to 2024"

### Time-Based Epidemiological Trends:

***Incidence Rate:***
IR = (New cases / Population at risk) × 100,000

***Adjusted Relative Risk:***
RR = (Incidence in exposed / Incidence in unexposed)
95% CI = exp[ln(RR) ± 1.96 × √(1/a + 1/m + 1/b + 1/n)]

### Geographic Distribution Analysis:

***Regional Variation Coefficient:***
RVC = σ/μ
Where: σ = √[Σ(xi - μ)²/n]

***Population-adjusted Rate Ratio:***
RR = (Cases_region/Population_region) / (Cases_reference/Population_reference)
95% CI = exp[ln(RR) ± 1.96 × √(1/O + 1/E)]

### Treatment Pattern Analysis:

***Response Rate:***
Response Rate = (Number of responders / Total treated) × 100
95% CI = p ± 1.96 × √(p(1-p)/n)

***Success Rate Comparison:***
χ² test statistic = Σ((O - E)²/E)

### Comorbidity Analysis:

***Cumulative Incidence:***
CI = (New cases during follow-up / Population at risk) × 100
95% CI = CI ± 1.96 × √(CI(1-CI)/n)

***Hazard Ratio:***
HR = h₁(t)/h₀(t)
95% CI = exp[ln(HR) ± 1.96 × SE(ln(HR))]

### Gender and Racial Disparity Analysis:

***Standardized Morbidity Ratio:***
SMR = Observed cases / Expected cases
95% CI = SMR ± 1.96 × √(Observed cases)/Expected cases

### Treatment Outcomes Analysis:

***Adjusted Odds Ratio:***
OR = (a/b)/(c/d)
95% CI = exp[ln(OR) ± 1.96 × √(1/a + 1/b + 1/c + 1/d)]
